# Revisiting the High-Benefit Patient: Generic Machine Learning Inference for Intensive Blood Pressure Control

**DOI:** 10.64898/2026.06.26.26356708

**Authors:** Amir Habibdoust, Xing Song

**Affiliations:** Institute for Data Science and Informatics, University of Missouri-Columbia, Missouri, USA; Department of Biomedical Informatics, Biostatistics, and Medical Epidemiology, University of Missouri-Columbia, Missouri, USA

## Abstract

We applied the model-free GenericML framework using robust estimators (the Best Linear Predictor, Group Average Treatment Effects, and baseline profile classification) to reassess treatment effect heterogeneity (HTE) of intensive blood pressure (BP) using pooled data from two large-scale randomized control trials on BP controls. Among 10,712 participants with up to three years of follow-up, we estimated the differential risks of the primary cardiovascular outcome. Intensive BP control produced a modest average risk reduction (BLP β_1_ = −0.0136), but evidence for HTE was not statistically significant (BLP β_2_ p = 0.443; GATES top–bottom contrast p = 0.471). Although CLAN grouped participants with metabolically adverse profiles into the highest predicted-benefit stratum, their treatment response did not differ significantly from that of the lowest-benefit group. Overall, while machine learning can identify clusters of high-risk baseline phenotypes, group-level inference revealed no meaningful HTE, underscoring the need for caution when interpreting individualized treatment predictions.

## 1 Introduction

Clinical decisions are rarely made for an “average” patient. Although randomized trials typically report average treatment effects (ATEs), clinicians must decide whether a given patient is likely to benefit or be harmed by treatment. Intensive blood pressure (BP) control illustrates this tension. While lowering systolic BP can reduce major cardiovascular events, it may also increase treatment burden, adverse events, and polypharmacy^1,2^. The net benefit of intensive BP control may therefore differ across patients depending on age, kidney function, baseline BP, comorbidity burden, and medication use^3^. Understanding heterogeneous treatment effects (HTEs), how treatment benefits and harms vary across individuals, is essential for translating trial results into patient-centered care.

Traditional subgroup analyses are the conventional approach to evaluating heterogeneity, but they have well-known limitations. Historically, most clinical datasets were too small to examine heterogeneity of treatment effects beyond a few prespecified subgroup splits, but modern datasets increasingly make it feasible to estimate more individualized variation in effects. A key impediment is the risk of spurious heterogeneity. Investigators may iteratively search across many candidate subgroups and selectively report extreme findings, inflating false positives and producing results that do not replicate ^4,5^. Although straightforward and easy to use, traditional subgroup analysis therefore has important limitations, including an increased risk of false positives when many variables are considered and reduced power to detect true effects^6,7^. Moreover, subgroup results are often hard to translate into personalized decisions. For instance, if intensive BP control appears beneficial among patients with chronic kidney disease (CKD) but harmful among those without CKD, it is unclear how to treat a patient with borderline kidney function or mixed risk features (e.g., mild CKD plus prior CVD). While one could define increasingly granular groups using multiple covariates, this approach further intensifies multiple testing and heightens concerns about cherry-picking, motivating principled statistical and machine-learning approaches that can model nonlinearities and higher-order interactions while controlling overfitting and false discovery^7^. These challenges have motivated rapid growth in causal machine-learning (CausalML) approaches to estimate HTEs and approximate individualized treatment effects (ITEs).

Methods such as causal forests can capture nonlinear relationships and high-order interactions, enabling estimation of HTEs and ITEs in high-dimensional settings^3,7–9^, in particular, applications to investigate treatment-effect heterogeneity in BP management. For example, Inoue et al.(2023)^7^ used a causal random forest to estimate ITEs of intensive systolic blood pressure control. Their analysis demonstrated that a “high-benefit” strategy (i.e., treating individuals with positive estimated treatment effects) could outperform traditional risk-based strategies, suggesting that patients with the highest predicted cardiovascular risk are not always those who benefit most from treatment. While CausalML methods can generate ITE predictions, these predictions often lack valid statistical inference, raising the risk that apparent “high-benefit patients” reflect model overfitting rather than true causal heterogeneity. However, applying these tools to causal heterogeneity analysis raises important inferential challenges. While these algorithms often perform well for prediction, obtaining valid statistical inference for heterogeneous treatment effects is substantially more difficult. In high-dimensional settings, causal forest estimators may fail to consistently estimate the conditional average treatment effect (CATE) without strong assumptions, and standard inference procedures may not remain valid across a broad class of data-generating processes ^10^.

To address this challenge, Chernozhukov et al (2025)^10^ proposed the Generic Machine Learning (GenericML) framework, which separates flexible prediction of treatment effects from the statistical inference on interpretable features of heterogeneity. Rather than attempting to estimate the full CATE function directly and risk overfitting, the framework focuses on inference for clinically interpretable summaries, such as the best linear projection (BLP), grouped average treatment effects (GATES), and classification analysis (CLAN), while using sample splitting and cross-fitting to ensure inference validity and unbiased quantification of uncertainty ^10^. GenericML provides a generic, model-free approach allowing the use of any machine-learning methods to predict and make statistically sound inferences on HTE.

In this study, we applied GenericML to re-evaluate a clinically and methodologically important question: can we robustly identify which patients benefit most from more intensive BP control? We pooled participant-level data from SPRINT and ACCORD-BP trials^1,2^ to estimate HTEs of intensive BP control on short-term cardiovascular outcomes via evaluating GATES across strata of predicted benefit and estimated a best linear projection (BLP) to quantify the magnitude of heterogeneity, and used classification analysis (CLAN) to characterize baseline profiles of higher-versus lower-benefit participants. By subjecting machine-learning predictions to strict inferential controls, we aim to demonstrate whether the data truly supports clinically actionable treatment effect heterogeneity.

## 2 Method

### 2.1 Data sources and study participants

We performed a pooled secondary analysis of participant-level data from two randomized control trials (RCTs) comparing intensive versus standard BP control: Systolic Blood Pressure Intervention Trial (SPRINT) and Action to Control Cardiovascular Risk in Diabetes Blood Pressure trial (ACCORD-BP) ^1,2^. Participant-level data for both trials were obtained from the National Heart, Lung, and Blood Institute (NHLBI) Biologic Specimen and Data Repository Information Coordinating Center (BioLINCC)^1^. Because this study involved the secondary analysis of fully anonymized, de-identified data, it was deemed exempt from full review by the Institutional Review Board at the University of Missouri (IRB # 2126482). SPRINT enrolled adults at elevated cardiovascular risk without diabetes, while ACCORD-BP enrolled adults with type 2 diabetes mellitus who were randomized within the BP intervention component. For the pooled dataset, we harmonized baseline covariates that were available in both trials and constructed trial indicators to preserve trial-level structure while enabling a combined analysis. The SPRINT trial demonstrated that targeting systolic BP <120 mmHg reduced major cardiovascular outcomes and mortality among high-risk adults without diabetes^1^. In contrast, ACCORD-BP ^2^ found no statistically significant reduction in the primary composite cardiovascular outcome among adults with type 2 diabetes in the overall trial population ^2^. (2). For the primary analysis, we included participants from both trials who: (a) had a valid assignment to either intensive or standard BP control; (b) had recorded baseline systolic and diastolic blood pressure measurements; and (c) had non-missing follow-up data required to ascertain the 3-year outcome. In sensitivity analyses, we relaxed criterion (c) and included all participants, including those lost to follow-up.

### 2.2 Intervention

For both trials, the intervention arm implemented intensive BP control targeting a systolic BP <120 mmHg, while the control arm followed the cohort-specific standard-of-care BP management. Randomization was inherited from the original RCTs, and we defined a binary treatment indicator, *D* (1 = intensive, 0 = standard), to represent arm assignment. The primary estimand was the difference in the 3-year risk of the primary composite cardiovascular outcome between the two arms.

### 2.3 Outcomes

The primary composite cardiovascular outcomes were harmonized across trials using each trial’s available primary composite event indicator and follow-up time. We set a fixed horizon *H* = 1,095 days (3 years). For each participant, we defined event incidence by 3 years as *Y*_3*y*_ = 1 (*event* = 1 & *time* ≤ *H*), and censoring before 3 years as *C*_<3*y*_ = 1(*event* = 0 & *time* < *H*). Participants with no event and follow-up time less than *H* were treated as censored prior to 3 years for the fixed-horizon analysis. The main analytic cohort excluded participants censored before 3 years (i.e., retained those with *C*_<3*y*_ = 0), and defined the outcome as *Y* = *Y*_3*y*_.

To enable pooled analysis, we harmonized a common set of baseline clinical indicators across trials using consistent variable names and units. Baseline covariates included demographics (age, sex), baseline blood pressure measurements (systolic blood pressure and diastolic blood pressure), adiposity (body mass index), kidney function measures (estimated glomerular filtration rate and serum creatinine), and lipid measures (total cholesterol, high-density lipoprotein cholesterol, and triglycerides). We also harmonized baseline medication indicators (statin use and aspirin use) and an antihypertensive treatment burden measure (number of antihypertensive medication classes at baseline). In addition, we harmonized comorbidity indicators: baseline cardiovascular disease history and chronic kidney disease status. Trial membership (SPRINT vs ACCORD-BP) and a diabetes trial indicator were retained to preserve trial context in pooled modeling. Race was available in SPRINT but was not harmonized for ACCORD in the current dataset and was therefore not used as a cross-trial covariate in pooled models.

### 2.4 Covariates

Baseline covariates were extracted at trial baseline and harmonized across trials. Variables included age, sex, baseline systolic BP and diastolic BP, body mass index, kidney function measures (eGFR, serum creatinine), lipid profile (total cholesterol, HDL, triglycerides), baseline antihypertensive medication count, statin use, aspirin use, prior cardiovascular disease status, and chronic kidney disease status, along with indicators for diabetes and trial membership.

### 2.5 Heterogeneous treatment effect estimation and inference using GenericML

The motivation of GenericML is that, although many machine-learning methods can generate useful predictors, uniformly valid inference for heterogeneous causal effects is difficult, and in high-dimensional settings ML methods may not consistently estimate conditional average treatment effect (CATE) without strong assumptions ^10^. GenericML addresses this challenge by shifting the inferential target from the full CATE function to interpretable features of heterogeneity that can be estimated with valid inference. Specifically, the framework proceeds in two stages.

In Stage 1, a chosen ML method (we used random forests) is trained to produce a *proxy* score of treatment effect heterogeneity, *S*(*Z*), where *Z* is a possible high-dimensional vector of covariates; importantly, GenericML does not require this proxy to be a consistent estimator of the true CATES ^10^. In Stage 2, the proxy is post-processed in a held-out (“main”) sample to estimate and conduct inference on three prespecified summaries. The Best Linear Predictor (BLP) of treatment effects using the ML proxy, Sorted Group Average Treatment Effects (GATES) across strata defined by the proxy, and Classification Analysis (CLAN) describing baseline characteristics of the most-vs least-benefit groups ^10^.

Specifically, let *s*_0_(*Z*) = *E*[*Y*(1) − *Y*(0) ∣ *Z*] denote the CATE, and let *S*(*Z*) be the cross-fitted ML proxy learned in Stage 1. The BLP is defined as:

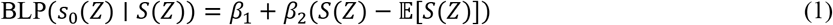

where (*β*_1_, *β*_2_)is the best linear projection of *s*_0_(*Z*)on *S*(*Z*). The GATE is defined as:

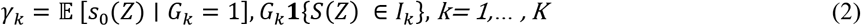

with *I*_*k*_ defined by prespecified quantile cutoffs of *S*(*Z*). In our implementation, GATE groups were defined using quintiles of the proxy S(Z), with cutoffs at 0.2, 0.4, 0.6, and 0.8. The CLAN summaries are:

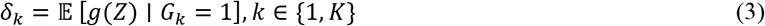

where *g*(*Z*)is a vector of baseline characteristics used to describe the lowest-vs highest-benefit groups.

GenericML uses sample splitting / cross-fitting and ensemble results across multiple random splits to reduce overfitting and ensure validity. We used 200 repeated sample splits and set α = 0.10 for inference. Inference is based on repeated splits into auxiliary and main samples, with quantile aggregation (e.g., medians of estimates and p-values; medians or quantiles of confidence interval endpoints) to stabilize conclusions and reduce split-induced variability. Point estimates and p-values were aggregated across splits using the median, and confidence intervals were aggregated using quantiles of split-specific interval endpoints. Analyses were run with a fixed random seed for reproducibility.

### 2.6 Sensitivity analyses

Expanding to all participants and accounting for loss to follow up, we conducted an inverse probability of censoring weighted (IPCW) ^11^ sensitivity analysis to evaluate the robustness of THE findings to differential follow-up patterns.. We modeled the probability of being observed through 3 years (i.e., *C*_<3*y*_ = 0) using separate logistic regression models within each trial, with baseline covariates as predictors. Weights were truncated to a prespecified range to limit the influence of extreme values, and participants with missing covariates required for the censoring model received missing weights and were excluded from the IPCW analysis. For the IPCW GenericML analysis, we constructed a weighted outcome using the event-by-3-years indicator, *Y*_*w*_ = *Y*_3*y*_ × *w*_IPCW_, and refit GenericML using the same covariate set and learners as in the main analysis. We compared BLP and GATES results between the main and IPCW analyses to evaluate the sensitivity of HTE conclusions to censoring adjustment.

## 3 Result

### 3.1 Cohort Pooling

The pooled analytic file included 4,733 ACCORD-BP participants and 9,361 SPRINT participants (total N = 14,094). Among them, 4,471/4,733 (94.5%) ACCORD-BP participants and 6,241/9,361 (66.7%) SPRINT participants were followed through 3 years, for a total of 10,712/14,094 (76.0%) included in the final analytic set. Early censoring before years occurred in 262 (5.5%) ACCORD-BP participants and 3,120 (33.3%) SPRINT participants. Among participants followed through 3 years (N = 10,712), treatment assignment remained balanced overall (standard: 5,358; intensive: 5,354) and within each trial (ACCORD-BP: 2,238 standard vs 2,233 intensive; SPRINT: 3,120 standard vs 3,121 intensive).

### 3.2 Baseline Characteristics

Among eligible participants, baseline characteristics were well balanced between the intensive and standard arms within each trial, consistent with randomization (Table 1). Across trials, participants in SPRINT were older than those in ACCORD (mean age approximately 67.5 vs 62.7 years) and had substantially lower kidney function (mean eGFR approximately 71.6 vs 91.5 mL/min/1.73m^2^), with a markedly higher prevalence of CKD (approximately 28.5% vs 8.5%). SPRINT participants also had lower statin use (approximately 44% vs 66%) and lower triglyceride levels (approximately 127 vs 194 mg/dL), whereas baseline SBP and DBP were similar across trials and treatment arms (SBP approximately 139 mmHg; DBP approximately 76-78 mmHg). In the pooled sample, most characteristics fell between the ACCORD and SPRINT trial-specific averages, indicating that the pooled cohort represents a mixture of two clinically different populations; this between-trial case-mix difference is important to consider when interpreting pooled heterogeneity analyses.

**Table 1.**
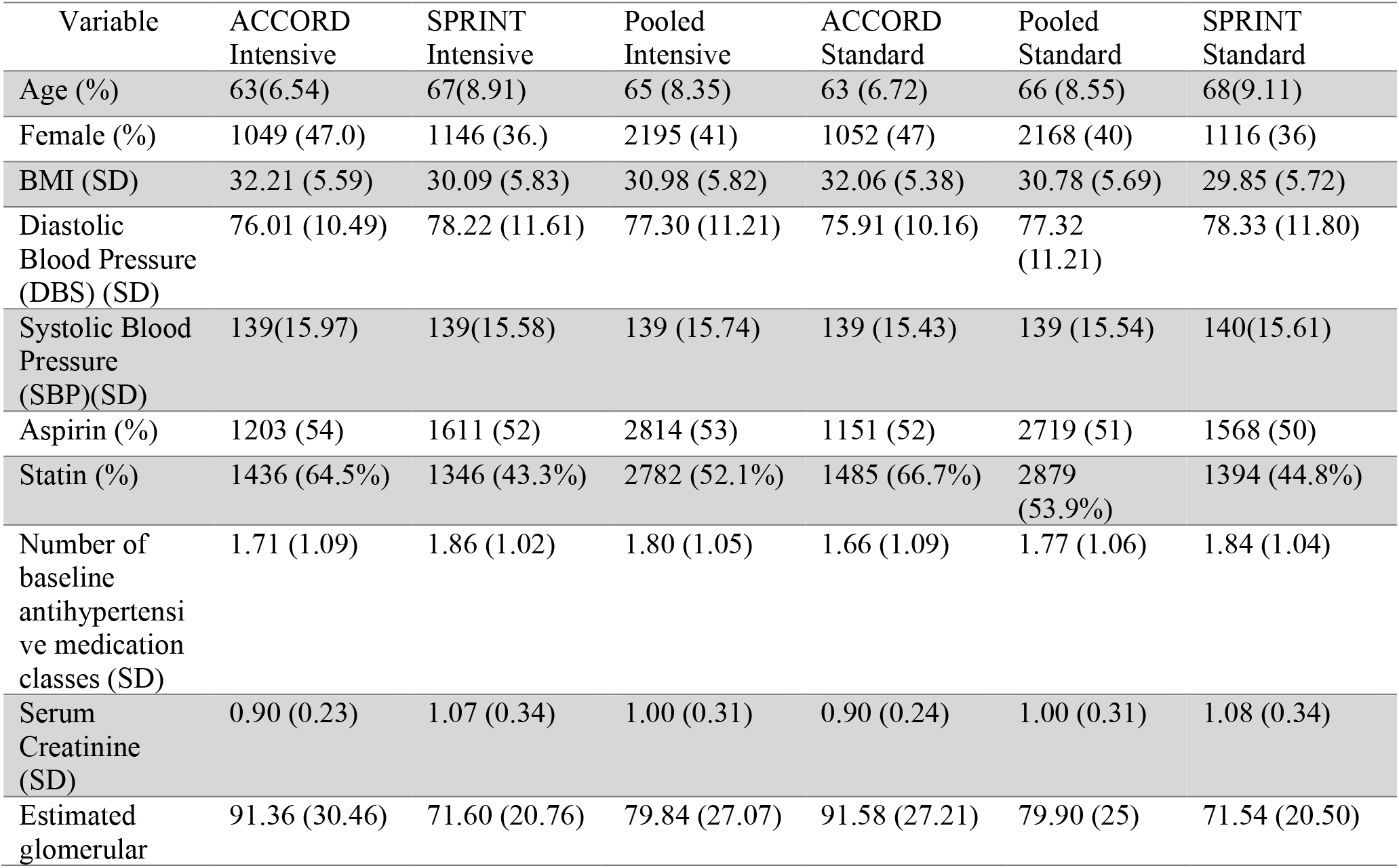
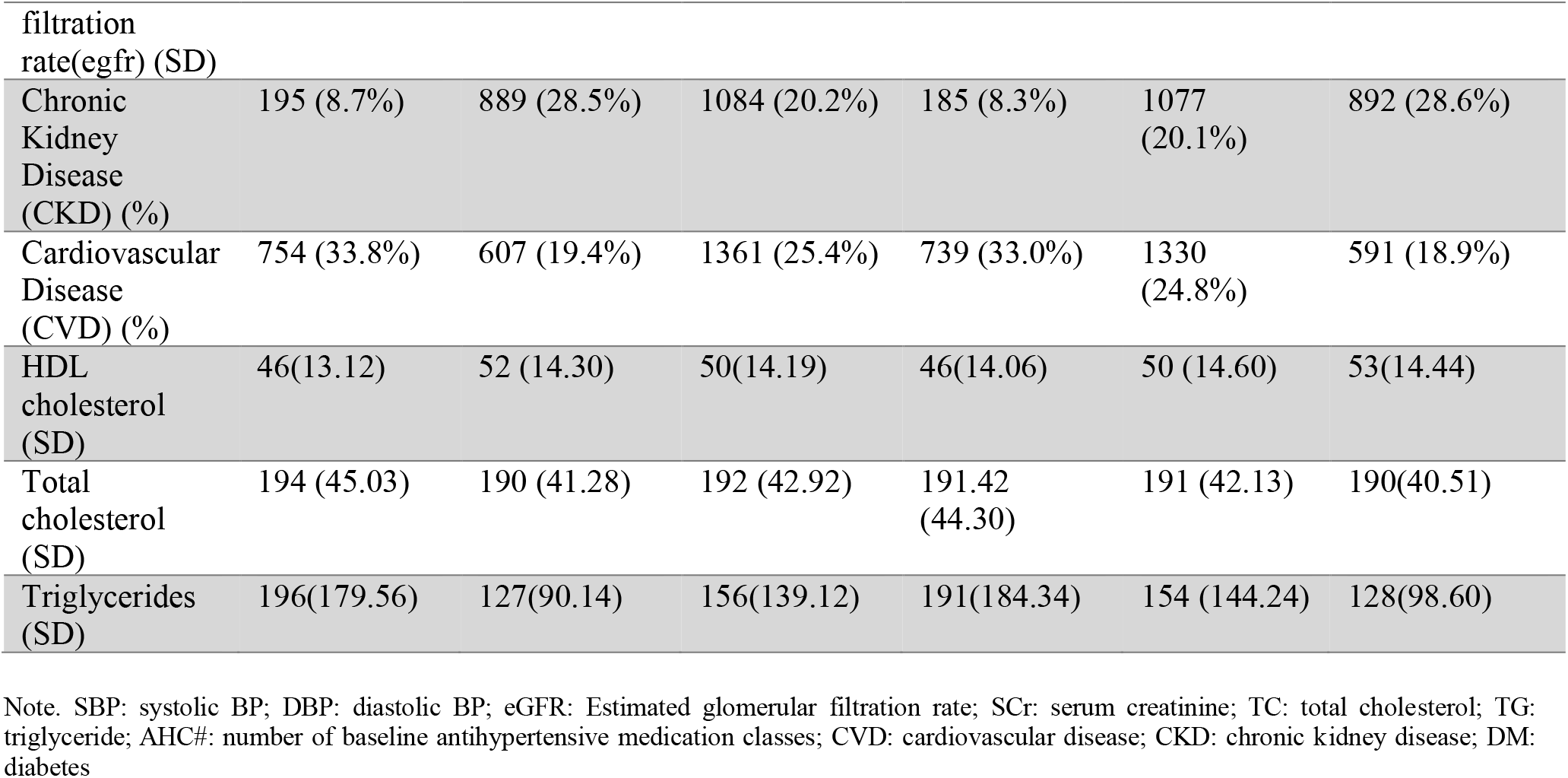
Baseline Characteristics.

### 3.3 Outcome Rates

The 3-year primary cardiovascular composite event risk differed by trial. The incidence rate was 0.0662 in ACCORD-BP and 0.0790 in SPRINT (Table 2). When pooling across trials, the 3-year event rate was 0.0799 in the standard arm and 0.0674 in the intensive arm. Stratified by trial and arm, event rates were 0.0652 (ACCORD-BP standard), 0.0672 (ACCORD-BP intensive), 0.0904 (SPRINT standard), and 0.0676 (SPRINT intensive).

**Table 2.** Best Linear Predictor (BLP)

| estimand | estimate |
| --- | --- |
| $\beta_1$ | $-0.0136$ CI $(-0.0275, 0.0002)$<br>[0.0536] |
| $\beta_2$ | $-0.0915$ CI $(-0.3192, 0.1334)$<br>[0.4432] |

Notes: Medians over 200 splits. Median confidence interval (alpha= 0.10) in parenthesis. P-values for the hypothesis that the parameter is equal to zero against the two-sided alternative in brackets.

### 3.4 Heterogeneous Treatment Effect Estimation: BLP, GATES

We found evidence of a modest average benefit of intensive blood pressure treatment, with limited evidence of effect heterogeneity. The Best Linear Predictor (BLP) indicated an average risk difference of −0.0136 (β_1_; 90% CI −0.0275 to 0.0002; p=0.0536). The heterogeneity slope (β_2_) was not statistically distinguishable from zero (−0.0915; 90% CI −0.3192 to 0.1334; p=0.443), suggesting no strong monotonic relationship between the learned HTE score and treatment effect.

Consistent with the BLP, Group Average Treatment Effects (GATES) (Table 3) estimated across five HTE-score groups were uniformly small and largely overlapping, with 90% confidence intervals that all crossed zero. The contrast between the highest- and lowest-score groups (*γ*_5_ − *γ*_1_) was −0.0160 (90% CI: −0.0595 to 0.0278; p=0.471), providing no evidence of meaningful separation between high- and low-benefit strata. Overall, these results suggest that while intensive treatment may reduce 3-year event risk on average, the data do not support clinically large heterogeneity across the derived HTE score distribution in the primary analysis.

**Table 3.**
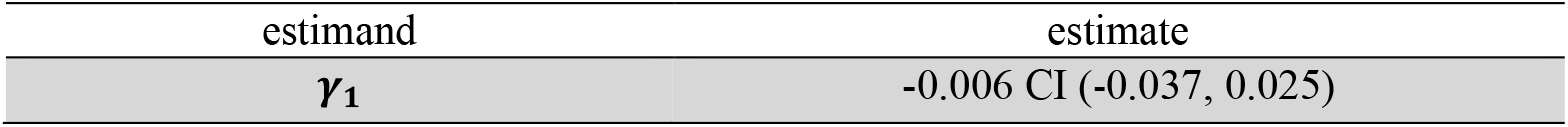
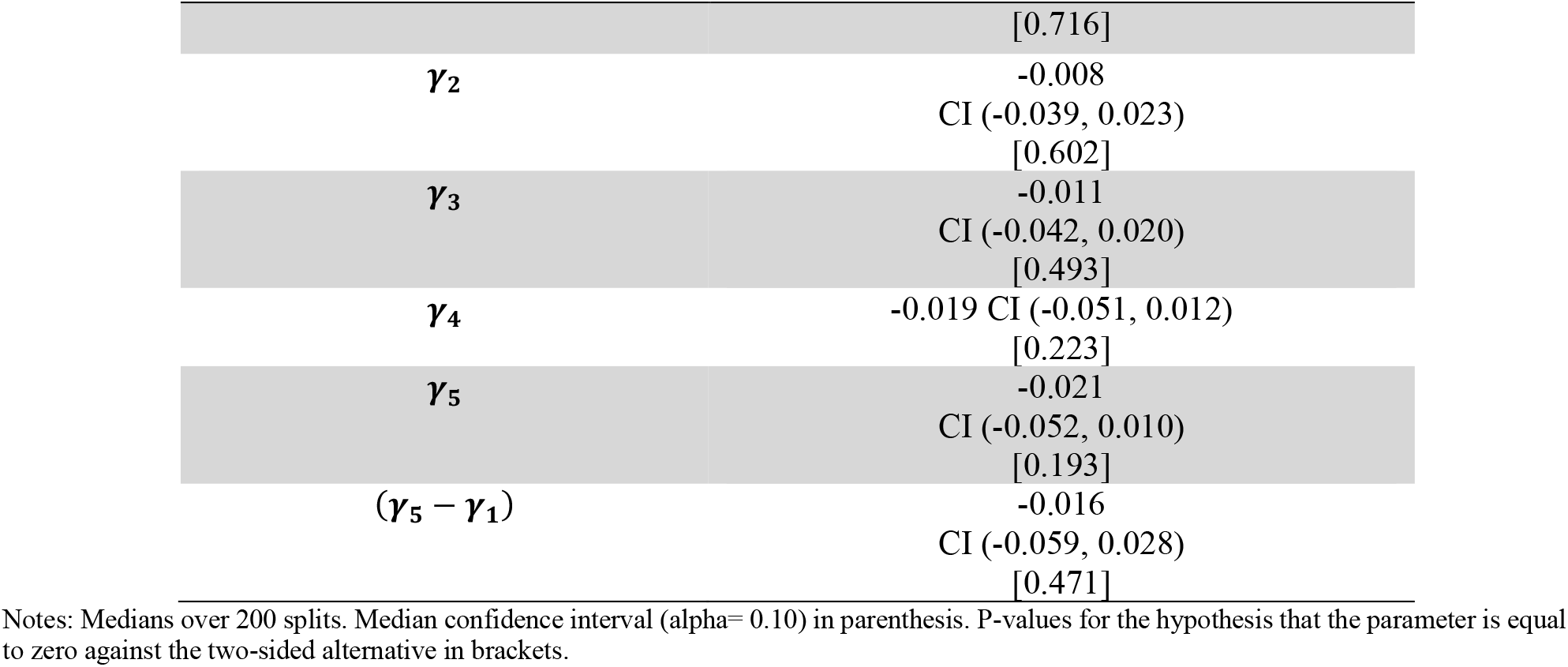
Group Average Treatment Effects (GATES)

**Table 4.**
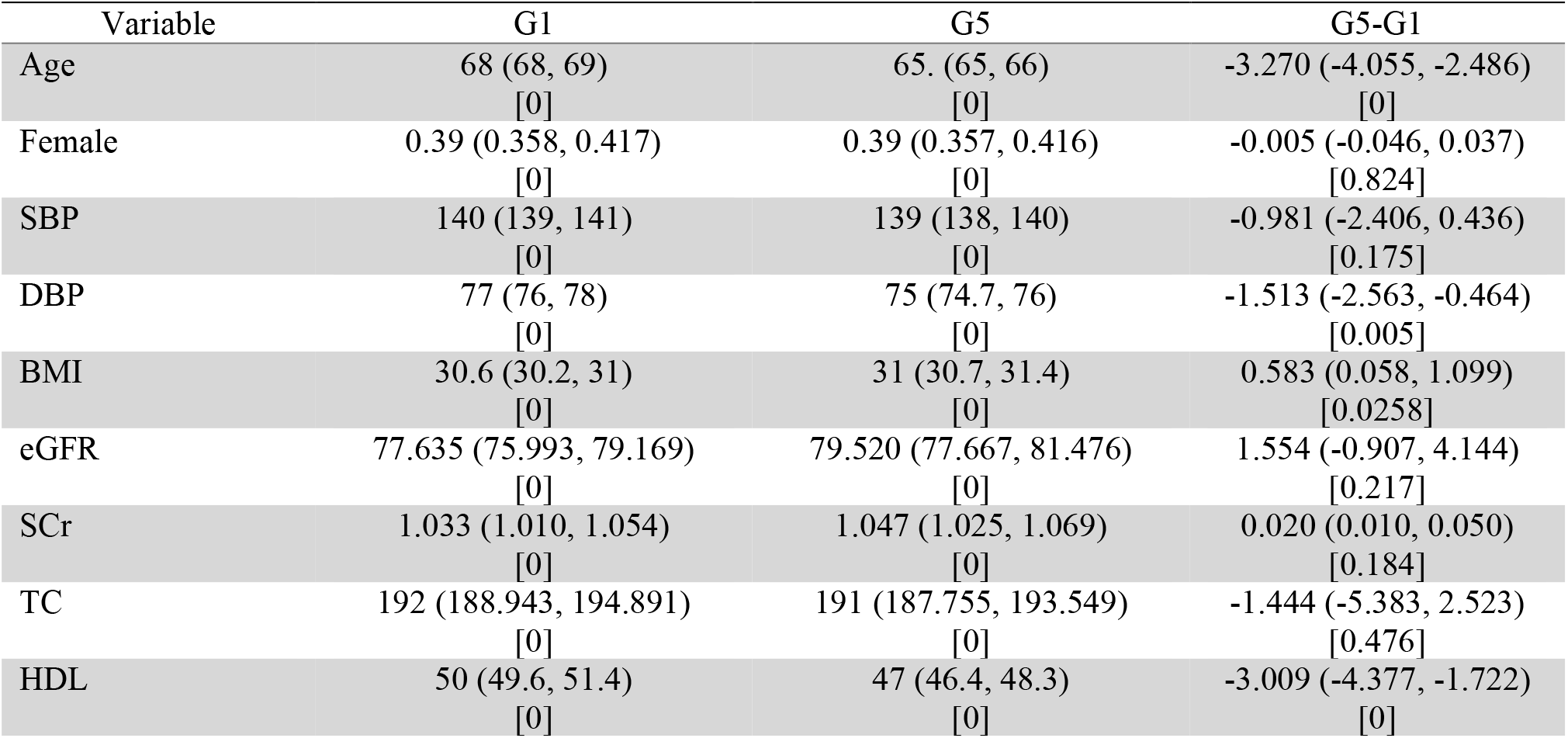
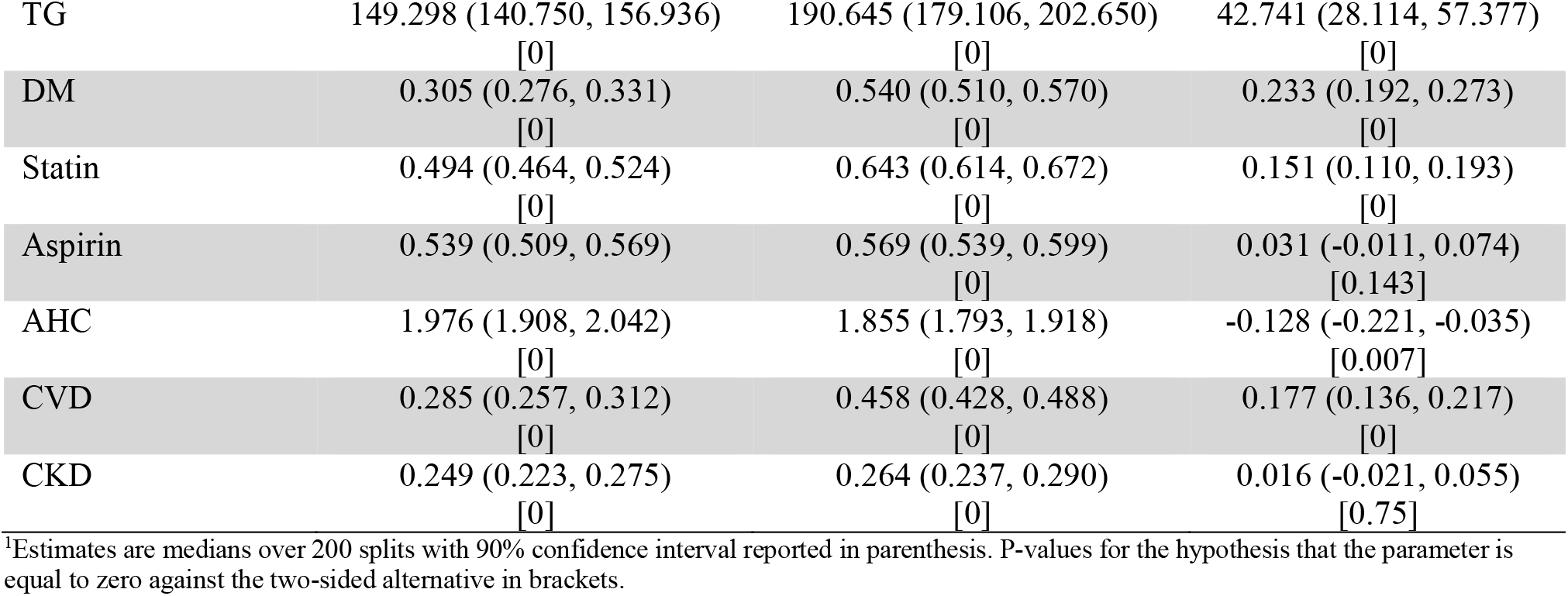
Classification Analysis (CLAN)

### 3.5 Characterizing High-benefit vs Low-benefit Groups: CLAN

In the CLAN analysis, we compared baseline characteristics between the lowest predicted benefit group (G1) and the highest predicted benefit group (G5). Several clinically meaningful differences emerged. Compared with G1, participants in G5 were younger (mean difference G5 - G1 = −3.27 years, 90% CI −4.06 to −2.49; p<0.001) and had slightly lower diastolic BP (−1.51 mmHg, 90% CI −2.56 to −0.46; p = 0.0047), while systolic BP did not differ significantly (−0.98 mmHg, 90% CI −2.41 to 0.44; p = 0.175). Metabolic and treatment-related profiles differed more strongly. G5 had higher BMI (+0.58 kg/m^2^, 90% CI 0.06 to 1.10; p = 0.026), lower HDL cholesterol (−3.01 mg/dL, 90% CI −4.38 to −1.72; p<0.001), and substantially higher triglycerides (+42.74 mg/dL, 90% CI 28.11 to 57.38; p<0.001), with no significant differences in total cholesterol, serum creatinine, or eGFR. G5 also showed higher prevalence of several clinical risk markers and treatments: statin use was higher (+0.151, 90% CI 0.110 to 0.193; p<0.001) and prior cardiovascular disease was markedly higher (+0.177, 90% CI 0.136 to 0.217; p<0.001). Diabetes prevalence was also substantially higher in G5 (+0.233, 90% CI 0.192 to 0.273; p<0.001). In contrast, female sex, aspirin use, and chronic kidney disease status did not differ significantly between groups (all p > 0.10). Finally, the number of baseline anti-hypertensive medication classes was slightly lower in G5 (−0.128, 90% CI −0.221 to −0.035; p = 0.007).

### 3.6 Sensitivity analysis: censoring adjustment (IPCW)

Under IPCW, the BLP results were qualitatively similar to the main analysis. The average effect estimate remained negative and of comparable magnitude (β_1_ = −0.0106; 90% CI −0.0215 to 0.00026; p = 0.0559), while the heterogeneity slope was not statistically significant (β_2_ = −0.0586; 90% CI −0.266 to 0.146; p = 0.562)(Table 5).

**Table 5.** Best Linear Predictor (BLP) - Sensitivity Analysis.

| Table 5- Best Linear Predictor (BLP) - Sensitivity Analysis |  |
| --- | --- |
| estimand | estimate |
| $\beta_1$ | -0.011 CI (-0.022, 0.000)<br>[0.056] |
| $\beta_2$ | -0.059 CI (-0.266, 0.146)<br>[0.562] |

Notes: Medians over 200 splits. Median confidence interval ( $\alpha = 0.10$ ) in parenthesis. P-values for the hypothesis that the parameter is equal to zero against the two-sided alternative in brackets.

GATES estimates across HTE score groups likewise did not show a clear monotone pattern, and the top–bottom contrast was not statistically significant (γ_5_−γ_1_ = 0.00973; 90% CI −0.0246 to 0.0440; p = 0.578) (see Table 6). While the point estimate for the top–bottom contrast flipped signs between the main analysis (−0.0160) and the IPCW analysis (+0.00973), neither estimate was statistically significant. This fluctuation further underscores that there is no stable, monotonic separation between the high- and low-predicted-benefit strata. Overall, censoring adjustment reinforced the interpretation of the main findings: the data do not support the presence of clinically meaningful treatment-effect heterogeneity. Under the IPCW sensitivity analysis, CLAN showed that the highest– vs lowest-score groups differed strongly on multiple baseline characteristics. Compared with G1, participants in G5 were younger, had higher baseline SBP, and had a markedly more “metabolic” profile with higher BMI, much higher triglycerides, and much lower HDL; statin and aspirin use were also higher. G5 also had fewer baseline antihypertensive agents, substantially higher baseline CVD history prevalence, and a much higher prevalence of diabetes. In contrast, CKD status showed little separation between G5 and G1.

**Table 6.**
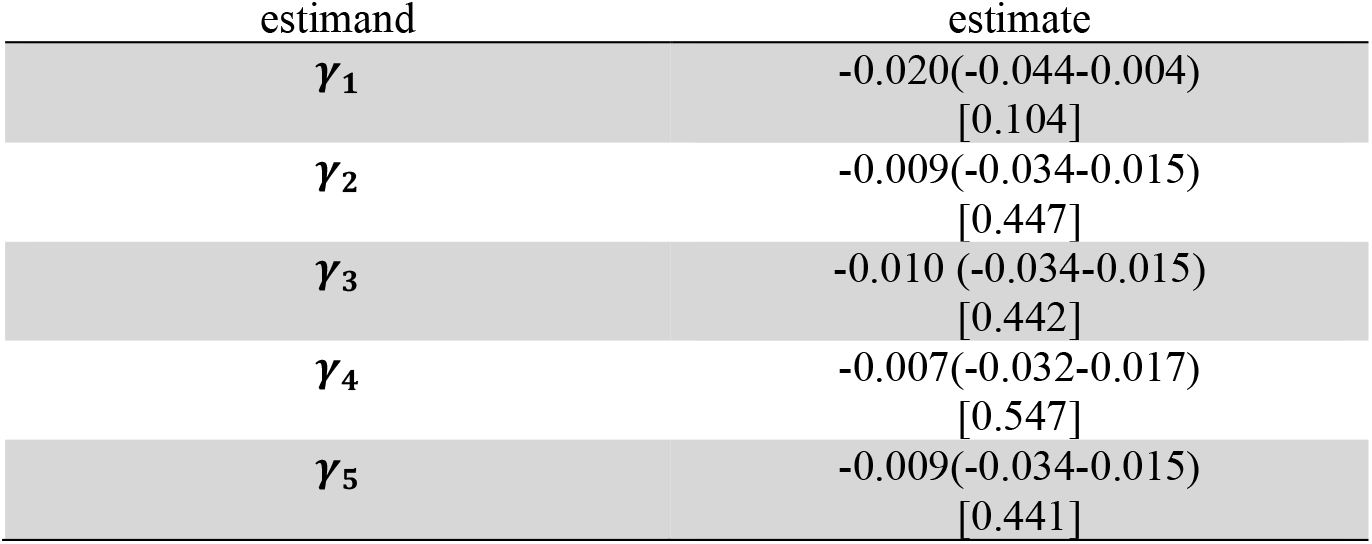
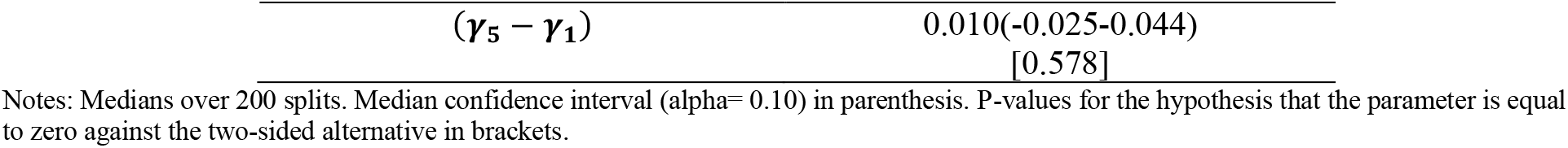
Group Average Treatment Effects (GATES)-Sensitivity Analysis.

In the IPCW sensitivity analysis, see Table 7, CLAN showed strong separation between the lowest predicted-benefit group (G1) and highest predicted-benefit group (G5). Compared with G1, G5 participants were younger (G5–G1 = −12.45 years; p<0.001) and slightly more often female (+0.040; p=0.024), with higher SBP (+2.03 mmHg; p=0.00058) and lower DBP (−1.62 mmHg; p=0.00020). G5 also had a more adverse cardiometabolic profile, including higher BMI (+3.28; p≈1×10^−54^), much lower HDL (−14.34; p<0.001), and much higher triglycerides (+92.57; p<0.001), along with slightly lower total cholesterol (−3.99; p=0.017). Comorbidity and treatment burden were greater in G5, with higher prevalence of statin use (+0.292; p<0.001), aspirin use (+0.058; p=0.0018), prior CVD (+0.301; p<0.001), and diabetes (+0.573; p<0.001), while the number of baseline antihypertensive medication classes was slightly lower (−0.382; p<0.001). Chronic kidney disease status did not differ significantly (p=0.304).

**Table 7.**
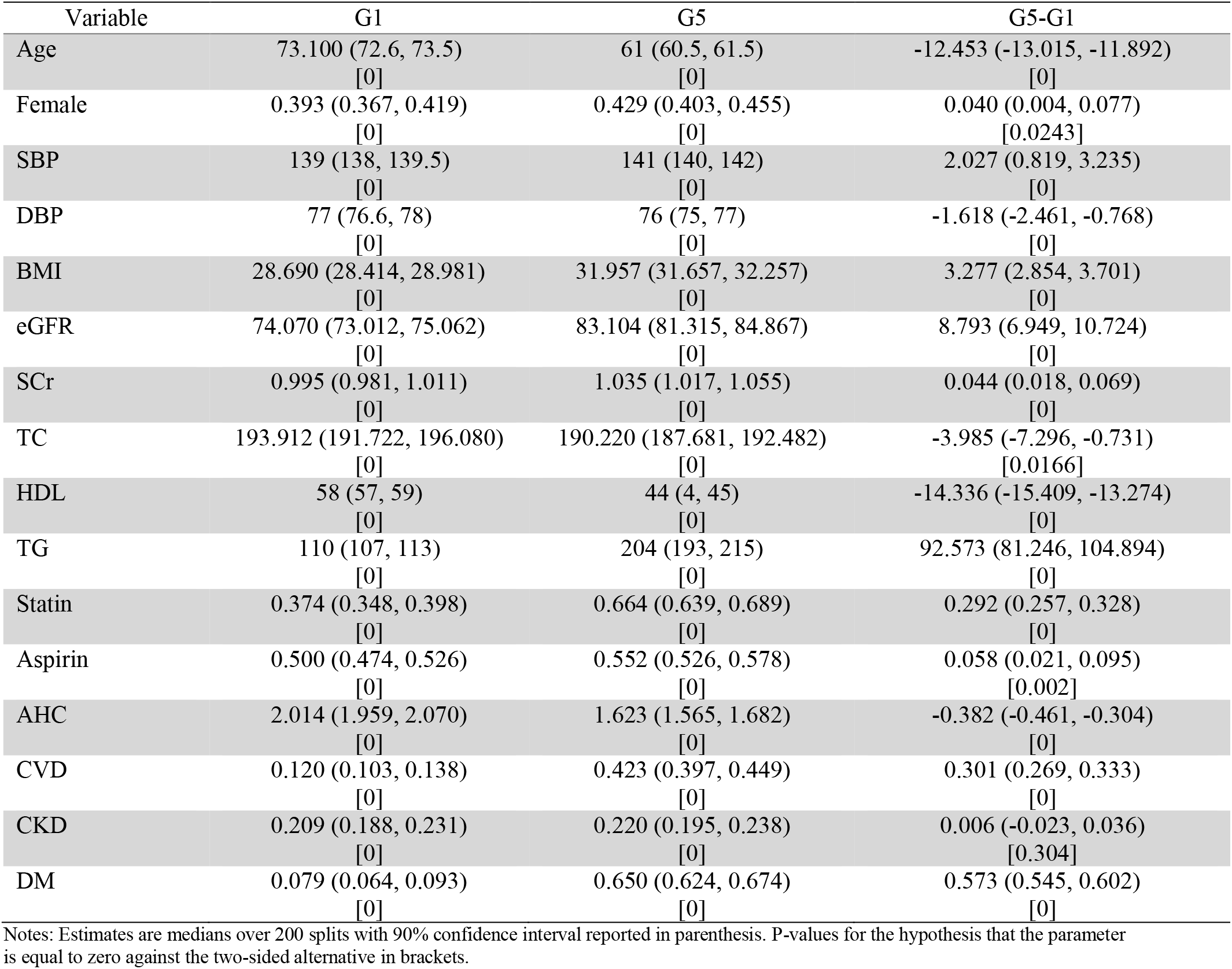
Classification Analysis (CLAN) - Sensitivity Analysis.

| Variable | G1 | G5 | G5-G1 |
| --- | --- | --- | --- |
| Age | 73.100 (72.6, 73.5)<br>[0] | 61 (60.5, 61.5)<br>[0] | -12.453 (-13.015, -11.892)<br>[0] |
| Female | 0.393 (0.367, 0.419)<br>[0] | 0.429 (0.403, 0.455)<br>[0] | 0.040 (0.004, 0.077)<br>[0.0243] |
| SBP | 139 (138, 139.5)<br>[0] | 141 (140, 142)<br>[0] | 2.027 (0.819, 3.235)<br>[0] |
| DBP | 77 (76.6, 78)<br>[0] | 76 (75, 77)<br>[0] | -1.618 (-2.461, -0.768)<br>[0] |
| BMI | 28.690 (28.414, 28.981)<br>[0] | 31.957 (31.657, 32.257)<br>[0] | 3.277 (2.854, 3.701)<br>[0] |
| eGFR | 74.070 (73.012, 75.062)<br>[0] | 83.104 (81.315, 84.867)<br>[0] | 8.793 (6.949, 10.724)<br>[0] |
| SCr | 0.995 (0.981, 1.011)<br>[0] | 1.035 (1.017, 1.055)<br>[0] | 0.044 (0.018, 0.069)<br>[0] |
| TC | 193.912 (191.722, 196.080)<br>[0] | 190.220 (187.681, 192.482)<br>[0] | -3.985 (-7.296, -0.731)<br>[0.0166] |
| HDL | 58 (57, 59)<br>[0] | 44 (4, 45)<br>[0] | -14.336 (-15.409, -13.274)<br>[0] |
| TG | 110 (107, 113)<br>[0] | 204 (193, 215)<br>[0] | 92.573 (81.246, 104.894)<br>[0] |
| Statin | 0.374 (0.348, 0.398)<br>[0] | 0.664 (0.639, 0.689)<br>[0] | 0.292 (0.257, 0.328)<br>[0] |
| Aspirin | 0.500 (0.474, 0.526)<br>[0] | 0.552 (0.526, 0.578)<br>[0] | 0.058 (0.021, 0.095)<br>[0.002] |
| AHC | 2.014 (1.959, 2.070)<br>[0] | 1.623 (1.565, 1.682)<br>[0] | -0.382 (-0.461, -0.304)<br>[0] |
| CVD | 0.120 (0.103, 0.138)<br>[0] | 0.423 (0.397, 0.449)<br>[0] | 0.301 (0.269, 0.333)<br>[0] |
| CKD | 0.209 (0.188, 0.231)<br>[0] | 0.220 (0.195, 0.238)<br>[0] | 0.006 (-0.023, 0.036)<br>[0.304] |
| DM | 0.079 (0.064, 0.093)<br>[0] | 0.650 (0.624, 0.674)<br>[0] | 0.573 (0.545, 0.602)<br>[0] |

Notes: Estimates are medians over 200 splits with 90% confidence interval reported in parenthesis. P-values for the hypothesis that the parameter is equal to zero against the two-sided alternative in brackets.

## 4 Discussion

In this pooled analysis of SPRINT and ACCORD-BP, we used the model-free GenericML framework to assess whether intensive BP control produces clinically meaningful heterogeneity in short-term cardiovascular risks. In the primary analysis, we found evidence consistent with a modest average benefit of intensive treatment (β_1_ ≈ −0.014), but limited evidence of significant heterogeneity along the learned HTE score. The BLP heterogeneity slope (β_2_) was not statistically distinguishable from zero, and GATES estimates across predicted-benefit strata were small with overlapping confidence intervals; the top–bottom contrast (γ_5_−γ_1_) was not significant. Overall, these results suggest that while intensive BP control may modestly reduce 3-year cardiovascular event risk on average, the data do not support large, monotone treatment-effect variation across the derived heterogeneity score in the primary analysis. In other words, we did not observe statistically strong evidence on generalizations of “high-benefit” or “low-benefit” subgroups.

CLAN summaries further characterized the baseline profiles underlying the learned heterogeneity score, providing insight into which clinical features were associated with higher versus lower predicted benefit from intensive BP control. In the primary analysis, participants in the highest-score stratum differed from those in the lowest-score stratum across several cardiometabolic and comorbidity indicators, including triglyceride levels, HDL cholesterol, statin use, diabetes status, and prior cardiovascular disease. These contrasts were even more pronounced in the IPCW sensitivity analysis, suggesting that accounting for potential informative censoring accentuated differences in baseline risk profiles across the score-defined groups. Importantly, CLAN is designed to describe who populates the high-versus low-score strata rather than to establish causal subgroup effects. The absence of clear separation in GATES estimates indicates that although these strata reflect clinically distinct baseline profiles, they did not correspond to clearly differentiated group-average treatment effects under the fixed 3-year risk-difference estimand.

Motivated by the study by Inoue et al. ^3^, who used a causal random forest to estimate ITEs and suggested that a “high-benefit” strategy can outperform conventional risk-based approach, we applied the GenericML framework to further evaluate the statistical validity of such “high-benefit” subgroup. Our analysis indicated only a modest and statistically non-significant increase in treatment benefit along the predicted HTE spectrum. Several methodological differences may account for this discrepancy. In particular, Inoue et al.^3^ focused on estimating individualized treatment effects and evaluating an ITE-based treatment rule, whereas GenericML evaluates prespecified group-level summaries of heterogeneity, such as BLP and GATES, using repeated sample splitting and cross-fitting. In addition, our pooled analysis relied on a harmonized covariate set available across both trials, which was narrower than the broader sociodemographic and clinical feature set used by Inoue et al.^3^, and the two studies also differed in their handling of missing data.

A key practical challenge in this pooled setting was the markedly different follow-up structure across trials under a fixed 3-year horizon, with substantially higher early censoring in SPRINT than ACCORD-BP. To address potential bias from differential follow-up, we performed an IPCW sensitivity analysis using trial-specific censoring models and truncated weights. IPCW results were qualitatively similar to the main analysis. The average effect remained modestly protective, β_2_ remained non-significant, and the GATES top–bottom contrast again showed no meaningful separation. Thus, censoring adjustment did not materially change the primary conclusion of limited heterogeneity.

This study has limitations. First, the fixed-horizon approach and restriction to observed 3-year status may introduce selection if censoring is informative; IPCW mitigates but cannot remove bias from unmeasured predictors of follow-up. Second, pooling trials with different eligibility criteria and baseline risk profiles may induce heterogeneity related to trial membership rather than biologic effect modification. Third, covariate harmonization was constrained by data availability (e.g., race was not harmonized across trials), and complete-case steps may affect generalizability.

## 5 Conclusion

While intensive BP control modestly reduces short-term cardiovascular risk on average, we found no rigorous evidence of clinically actionable treatment-effect heterogeneity. Although our GeneriML approach successfully clustered participants with a highly metabolic phenotype into a predicted “high-benefit” group, this subgroup did not demonstrate a statistically confirmed treatment advantage compared with the “low-benefit” group. These findings challenged prior causal forest analyses, warranting the importance of requiring strict inferential frameworks to guard against the over-interpretation of spurious heterogeneity when translating machine learning predictions into personalized clinical guidelines.

## Data Availability

The participant-level data analyzed in this study were obtained from the National Heart, Lung, and Blood Institute Biologic Specimen and Data Repository Information Coordinating Center (BioLINCC). Access to the SPRINT and ACCORD-BP data is available to qualified researchers through BioLINCC upon submission and approval of a data request. The analytic code and derived non-identifiable results may be made available from the corresponding author upon reasonable request, subject to applicable data-use agreements.

## Footnotes

1 Data are available through National Heart, Lung, and Blood Institute BioLINCC data repository at https://biolincc.nhlbi.nih.gov/studies/sprint/ and https://biolincc.nhlbi.nih.gov/studies/accord/

## Notes

### Competing Interest Statement

The authors have declared no competing interest.

### Author Declarations

The Institutional Review Board of the University of Missouri waived ethical approval for this work because it involved a secondary analysis of fully de-identified participant-level data from the SPRINT and ACCORD-BP randomized trials (IRB #2126482).

### Summary of Updates

The coauthor's name has been corrected from Xing Sing to Xing Song.

